# Ankle proprioception and the relationship to rigidity in Parkinson’s disease

**DOI:** 10.1101/2024.08.16.24312075

**Authors:** Jacquelyn V.L. Sertic, Jason Kang, Colum D. MacKinnon, Jürgen Konczak

## Abstract

**Introduction:** Proprioceptive mechanoreceptor afferents form the basis for the perception of body position and muscle tone regulation. Parkinson’s disease (PD) is associated with proprioceptive dysfunction. Proprioceptive dysfunction at the ankle impairs balance and gait. No comprehensive data exist characterizing the extent of ankle position sense dysfunction in PD and its relationship with abnormal muscle tone.

**Objectives:** (1) Quantify ankle position sense acuity in PD. (2) Determine the relationship between ankle position sense and muscle rigidity.

**Methods:** Sixteen people with mild to moderate PD and 16 age-matched controls participated. PD participants were OFF medication during testing. Using a 2-forced-choice psychophysical paradigm, a participant’s ankle was passively rotated to two distinct positions – a reference of 15° plantarflexion and a smaller amplitude comparison position. Subsequently, the user verbally indicated which position was perceived as more plantarflexed. From the stimulus-response data a just-noticeable-difference (JND) threshold and uncertainty area (UA) were derived as measures of ankle position sense acuity. Muscle rigidity was assessed using the MDS-UPDRS III scale (item 3.3).

**Results:** Both median JND threshold (PD: 2.1°, controls: 1.5°) and UA (PD: 1.7°, controls: 1.1°) were significantly elevated in the PD group compared to controls (*p* < 0.05). Seven participants with PD (44%) had thresholds above the control group maximum. JND threshold correlated positively with lower extremity rigidity (ρ = 0.50, *p =* 0.047).

**Conclusion:** PD can lead to reduced ankle position sense acuity. Lower proprioceptive acuity tended to be associated with higher rigidity suggesting that PD broadly alters proprioceptive signal processing affecting both perception and regulation of muscle tone.

## INTRODUCTION

Parkinson’s disease (PD) is a progressive neurodegenerative disease that alters the neural processing within somatosensory and motor networks. As a result, people with PD exhibit a range of somatosensory and motor symptoms, such as impaired limb proprioception, tremor, bradykinesia, and increased muscle rigidity (Goetz, 2011; Konczak et al., 2009). It is known that proprioceptive mechanoreceptor afferents from the lower limbs are essential for the control of balance and gait and the regulation of leg muscle tone (Takakusaki, 2017). Muscle tone is regulated through a task-dependent modulation of stretch reflexes, which rely on feedback from proprioceptive mechanoreceptors. There is evidence that proprioceptive mechanoreceptors are unaffected, but long-latency stretch reflexes are abnormal, in people with PD (Tatton & Lee, 1975). This supports the notion that the degraded proprioceptive acuity and elevated muscle tone arise from impaired central processing of proprioceptive afferents and alterations to fusimotor drive which can affect the sensitivity of the muscle spindles (Asci et al., 2023; Lee & Tatton, 1975; Seiss et al., 2003; Tatton & Lee, 1975).

There are numerous reports indicating impaired proprioceptive perception in the upper extremities of people with PD (Konczak et al., 2007, 2008, 2012; Maschke et al., 2003, 2006; Putzki et al., 2006). Based on these results, an altered proprioceptive body map due to PD has been suggested, assuming similar proprioceptive deficits in the lower extremities (Konczak et al., 2009). Yet, current empirical evidence of lower limb proprioceptive dysfunction in PD is inconclusive. Higher position matching errors at the knee (Ribeiro Artigas et al., 2016) and for ankle inversion (Teasdale et al., 2017) were reported for people with PD in comparison to healthy controls. However, because the applied methods required participants to make an active voluntary movement, it becomes impossible to dissociate the known motor dysfunction in PD from the assumed dysfunction in proprioceptive perception (e.g., position sense). Rather, the derived error measures are indicative of neural processes underlying proprioceptive-motor integration. Finally, there is an incomplete understanding of how deficits of proprioceptive perception and muscle tone regulation are related in PD.

The present study sought to close these knowledge gaps. We aimed (1) to assess ankle position sense by applying a psychometric method that does not rely on active movement such that the outcome measures are not contaminated by motor control processes, and (2) to relate the psychometric measures of ankle proprioceptive perception to an established clinical measure of lower limb rigidity.

## METHODS

### Participants

Sixteen participants with idiopathic PD and sixteen age-matched controls volunteered for this study. Healthy controls were age-matched within 3 years of a PD participant. For relevant demographics of study participants, see **Table 1**. Inclusion criteria were: (1) Age between 18-95 for participants of both groups, (2) confirmed diagnosis of idiopathic PD. Exclusion criteria were: (1) Inability to consent as assessed by the UCSD Brief Assessment of Capacity to Consent score <14, (2) clinical diagnosis of peripheral neurologic pathology, (3) deep brain stimulation or other neurosurgery, (4) tremor larger than 1 cm in the OFF-medication state, (5) inability to achieve at least 24° of ankle range of motion, (6) exposure to chemotherapy, (7) previous or current use of benzodiazepine, (8) lower extremity orthopedic or musculoskeletal injury within the last six months, (9) lower limb amputation, knee replacement, or presence of lower limb pain, and (10) participants with PD were excluded if they had any neurological disorder other than idiopathic PD. Healthy controls did not have any diagnosis of neurological conditions. To determine eligibility prior to participating, all participants provided a verbal medical history using REDCap electronic data capture tools hosted at the University of Minnesota (Harris et al., 2009, 2019; Lawrence et al., 2020). Cognitive function was assessed using the Montreal Cognitive Assessment (MOCA) (Nasreddine et al., 2005). Clinical characteristics of participants with PD are summarized in **Table 2**.

**Table 1.** Demographics of study participants.

| | **Healthy controls**<br>(n = 16)<br>mean ± SD (range) | **Parkinson's disease**<br>(n = 16)<br>mean ± SD (range) |
| --- | --- | --- |
| Female / Male | 7 / 9 | 10 / 6 |
| Age (years) | 66.7 ± 6.4 (52.4 – 75.6) | 66.9 ± 6.3 (55.1 – 75.3) |
| Disease Duration<br>(years) | NA | 6.6 ± 3.0 (0.5 – 11.7) |
| MDS-UPDRS III<br>Total Score<br>*OFF medication* | 3 ± 1 (0 – 4) | 31 ± 12 (15 – 52) |
| MDS-UPDRS III<br>Section 3.3<br>total rigidity score | 0 ± 1 (0 – 2) | 7 ± 4 (1 – 14) |
| Rigidity score<br>of more affected<br>limb | 0 ± 0.3 (0 – 1) | 1 ± 1 (0 – 3) |
| MOCA Score | 28 ± 2 (25 – 30) | 27 ± 2 (23 – 30) |
| PA Score | 47 ± 22 (9 – 83) | 61 ± 44 (25 – 185) |

MDS-UPDRS-III = Movement Disorders Society – Unified Parkinson's Disease Rating Scale; MOCA Score = Montreal Cognitive Assessment; PA score = Godin Shephard Activity Scores < 14 are Insufficiently Active, 15-23 are Moderately Active, and >24 are Active.

**Table 2.** Clinical characteristics of participants with Parkinson’s disease.

| **ID** | **Age range (years)** | **More affected side** | **Disease duration (years)** | **H&Y** | **MDS-UPDRS-III OFF state** | **MOCA** | **LED (mg/diem)** | **Medication** |
| --- | --- | --- | --- | --- | --- | --- | --- | --- |
| 1 | 70-75 | R | 9.8 | 2 | 24 | 26 | 900 | L IR |
| 2 | 70-75 | L | 11.7 | 3 | 39 | 28 | 750 | L IR & ER |
| 3 | 56-60 | R | 8 | 2 | 29 | 30 | 450 | L IR |
| 4 | 60-65 | R | 5.6 | 2 | 16 | 27 | 900 | L IR |
| 5 | 66-70 | R | 6.9 | 2 | 39 | 26 | 600 | L IR, THP |
| 6 | 60-65 | L | 4.8 | 2 | 34 | 26 | 600 | L IR |
| 7 | 70-75 | L | 6 | 2 | 16 | 30 | 1100 | L IR, PH |
| 8 | 70-75 | L | 4.2 | 2 | 48 | 29 | 750 | L IR |
| 9 | 50-55 | R | 6.9 | 2 | 19 | 30 | 600 | L IR |
| 10 | 66-70 | R | 11.7 | 2 | 31 | 29 | 1050 | L IR, AH, PH |
| 11 | 60-65 | R | 3.4 | 1 | 25 | 23 | 600 | L IR |
| 12 | 70-75 | L | 7.4 | 3 | 52 | 28 | 1131 | Rytary |
| 13 | 56-60 | L | 0.5 | 2 | 24 | 29 | 650 | L IR |
| 14 | 66-70 | R | 5.1 | 2 | 15 | 27 | 300 | Selegiline, oral |
| 15 | 60-65 | R | 5 | 2 | 42 | 23 | 700 | L IR |
| 16 | 70-75 | R | 8.4 | 2 | 38 | 26 | 750 | L IR & ER |

MDS-UPDRS-III = Movement Disorders Society – Unified Parkinson's Disease Rating Scale; Medication: L IR = Carbidopa/Levodopa Immediate Release, L ER = Carbidopa/Levodopa Extended Release, THP= Trihexyphenidyl, PH = pramipexole; Levodopa equivalent dose (LED) = 100 mg standard levodopa equals 75 mg extended release levodopa, 100 mg pramipexole, 60 mg rytary, 10 mg oral selegiline(Tomlinson et al., 2010)

Participants with PD were recruited from the University of Minnesota movement disorders outpatient clinic and an IRB-approved registry of former study participants. All participants provided written informed consent prior to participation in the study. The experimental protocol was approved by the University of Minnesota Institutional Review Board (STUDY00018992).

Participants with PD were in their clinically defined OFF medication state during testing. They abstained from taking their medication for 12 hours for immediate-release and extended-release medications that are taken more than one time per day. Extended-release medications taken only once per day were withdrawn for 24 hours prior to the start of the data collection.

### Testing Apparatus

Ankle position sense acuity was measured using the Ankle Proprioceptive Acuity System (APAS, **Figure 1A**). The APAS is a one-degree-of-freedom passive motion manipulandum that rotates the ankle in the sagittal plane (dorsiflexion-plantarflexion). Participants placed their foot on the platform at a neutral ankle joint position (approximately 90° with respect to the longitudinal axis of the shank). The foot platform was set to the height of the foot, such that the lateral malleolus was aligned to the center of rotation of the APAS. This allowed the APAS to rotate about the center of rotation of the ankle joint. A Velcro strap secured the distal portion of the foot to the foot platform during testing. Using the handle, an experimenter rotated the foot passively from the starting neutral foot position to other desired target positions. To ensure precise repeatable joint position, metal pins were inserted into a pegboard, restricting the system from moving past the starting and stopping positions. The smallest difference between ankle positions was 0.1°. Angular position was recorded by an optical encoder at a sampling frequency of 42.6Hz. Validity of the APAS system and its outcome measures was established previously (Mahnan et al., 2020).

**Figure 1.**
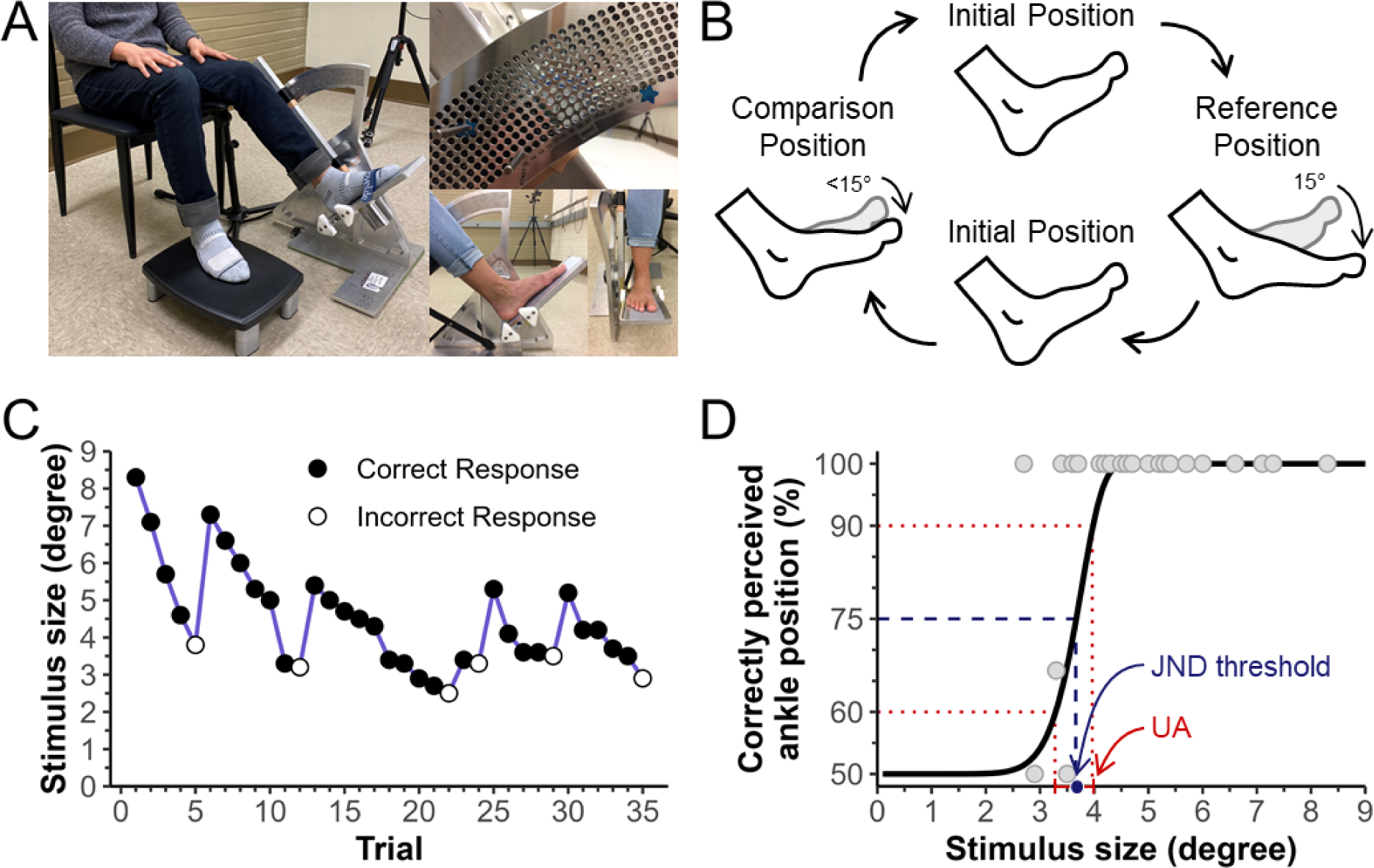
**(A)** Ankle position sense experimental set up. The examiner adjusts the footrest to align the ankle with the system center of rotation before starting the testing procedure. (Top Right) A subset of holes in the pegboard. Each hole in the vertical direction indicates a 0.1-degree difference in angle and in horizontal direction illustrates a 1-degree difference. Metal stoppers are inserted to a specific hole to identify the initial and target locations of ankle position. **(B)** Schematic of a single trial of ankle position sense testing. The foot is initially at a 0° position with the shank 90° from the longitudinal axis of the foot. **(C)** Exemplar stimulus size and response sequence of one participant. **(D)** Grey dots are the stimulus sizes from (C) at their correct response rate. The Just-Noticeable-Difference (JND) threshold is defined as the stimulus size at the 75% correct response rate. JND threshold is represented by the dashed blue line. Uncertainty area (UA) is calculated as the difference in stimulus sizes between 90% and 60% correct response rate. UA is represented as the range between the dotted red lines.

### Procedure

Prior to testing, motor symptom severity, including muscle rigidity, was evaluated by a single investigator (JVLS) using the MDS-UPDRS III rating scale. Position sense testing took place in a quiet room. Participants sat in an upright position. The more affected foot rested on the APAS platform, but the leg was unloaded (see **Figure 1A**). Participants wore vision occluding goggles.

Each trial consisted of passively rotating the ankle to two target positions (**Figure 1B**) at an angular velocity of 6.2 ± 1.0 deg/s. The experimenter would rotate the ankle from the initial neutral position (0°) to the reference position, which was always 15° of plantarflexion and held this position for two seconds. Then the foot would be returned to the neutral position. Subsequently, the foot was rotated to the comparison position (<15° plantarflexion), held for two seconds and then returned to neutral. Presentation of the reference and comparison positions within a trial (first or second) was randomized between trials. We applied a 2-forced-choice psychophysical paradigm requiring participants to verbally indicate which position, the first or the second, they perceived to be more plantarflexed. Knowledge of results was never provided to the participant to exclude possible learning effects. After each trial, the comparison position for the next trial was determined using the Bayesian inference-based adaptive psi-marginal algorithm. The algorithm identified the next comparison position using angular difference between the reference and the prior comparison position and the correctness of the participant’s prior responses (see **Figure 1C**) (Prins, 2013). Each test comprised 35 trials. Breaks were taken every seven trials to mitigate fatigue. During testing, participants did not wear shoes, but could choose to wear socks.

To ensure that the participants did not actively move or activate involved muscles during ankle position sense acuity testing, electromyographic recordings of the tibialis anterior and medial and lateral heads of the gastrocnemius were recorded during each trial using the Delsys Trigno System DS-T03. Trials were repeated if muscle activity increased for longer than 250ms above their resting muscle activity level. Some patients with PD exhibited rigidity during testing. That is, muscle activity increased upon passive displacement without voluntary muscle activation. For those participants, muscle activity level during displacement was used as the baseline reference. Any muscle activity above that induced by the rigidity was noted as a voluntary activation and the trial was repeated.

### Outcome measures

After test completion, a log-Weibull function was fitted to the stimulus size difference and verbal response data of each participant (**Figure 1D**, Prins, 2013). Based on the fitted function, two measurements of ankle position sense acuity were calculated: (1) *Just-noticeable-difference (JND) threshold* and (2) *uncertainty area (UA)*. The JND threshold was the stimulus size difference at the 75% correct response rate. This is a measure of proprioceptive bias (i.e., systematic error) with a smaller JND threshold indicating high position sense acuity. The UA was calculated as the difference between the stimulus sizes at 60% and 90% correctly perceived ankle position. This is a measure of precision (i.e., random error). A smaller UA indicates more certainty when discriminating between two stimuli. A measure of lower limb rigidity was derived from the MDS-UPDRS item 3.3 lower extremity scores (Goetz et al., 2008). Only the score of the more affected leg is reported and was related to proprioceptive outcome measures, which were also obtained for the more affected limb for the participants with PD or the yoked limb for the controls.

### Data Analysis

The distribution of all variables was examined for normality using Shapiro-Wilk’s test of normality. Demographic features were compared between groups using independent samples t-test for Age and Mann-Whitney U tests for MOCA scores. Position sense JND threshold was normally distributed for both groups and uncertainty area was not normally distributed for the parkinsonian group. For consistency, non-parametric Mann-Whitney U tests were performed for both outcome measures to determine group differences. Spearman’s rank-order correlations were used to determine the relationships between lower extremity rigidity and other continuous outcome measures such as position sense acuity and disease duration. Pearson product-moment correlations were performed to determine relationships between position sense acuity and motor severity, duration of disease, dosage of medication, and cognitive function. Effect size was computed using rank-biserial correlation coefficient for the Mann-Whitney U tests. Outliers were identified as greater than 3 times the interquartile range. One outlier was detected and removed from analysis in the PD group for UA. The significance level for all tests was set to α = 0.05. All statistical analyses were conducted using R software version 4.3.1 (R Core Team, 2023).

## RESULTS

Age and MOCA scores were comparable between groups (see **Table 1**; *p* > 0.05).

### Ankle position sense acuity between groups

Six of the 16 participants with PD (38%) had JND thresholds larger than the maximum of the healthy controls (**Figure 2A**). In those PD participants with elevated thresholds, median JND threshold was 3.6°. In contrast, median JND threshold for those within the normal range was 1.8°. Group median JND thresholds were 1.5° (range: 1.0° – 2.7°) for controls and 2.1° (range: 1.2° – 4.4°) for the PD group, which yielded a statistically significant difference (*z* = 66, *p* = 0.020) with a moderate effect (*r* = 0.41). To appreciate these data, consider that the range of JND threshold for the Parkinsonian group was 1.2° – 4.4°, meaning that the smallest difference from 15° that could be perceived with 75% accuracy was 13.8° and the largest was 10.6°.

**Figure 2.**
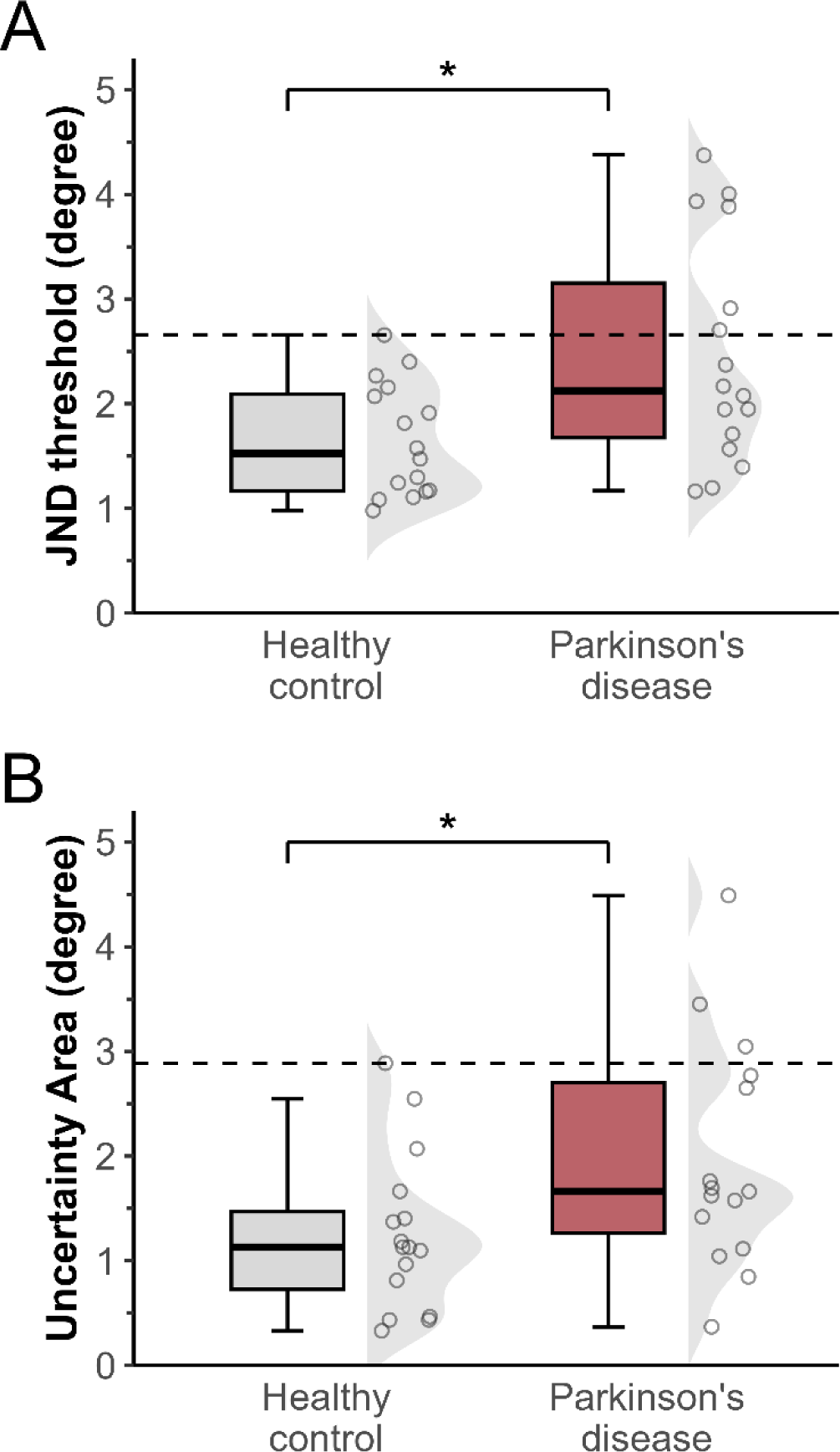
Ankle position sense acuity between people with Parkinson’s disease (PD) and healthy controls. **(A)** JND position sense threshold and **(B)** UA position sense values. The lower end of the box represents the 25^th^ percentile and the upper end of the box indicates the 75^th^ percentile. The line within the box is the median. The whiskers indicate 1.5 times the interquartile range of the respective distribution. Each datapoint represents an individual threshold. Both position sense acuity outcome measures were larger in the parkinsonian group than the control group (\**p* < 0.05).

**Figure 2B** illustrates the distribution of the Uncertainty Area values for both groups. One outlier was identified and removed from the Parkinsonian group. Three of the 15 (20%) participants with PD had UA values outside the range of controls. Median UA was 1.1° (range: 0.3° – 2.9°) for controls and 1.7° (range: 0.4° – 4.5°) for the PD group. The group medians were significantly different from each other (*z* = 68.5, *p* = 0.044) with a moderate effect (*r* = 0.37). Median UA for the Parkinsonian group was 1.7° meaning that for a JND threshold of 2.1°, participants could correctly perceive their ankle position 60% of the time at 1.3° and 90% of the time at 3°. Overall, 44% of individuals with PD exhibited impairments in either JND threshold, UA, or both.

### Relationship between ankle proprioception and rigidity

The distribution of the rigidity score is illustrated in **Figure 3A**. Fifteen controls demonstrated rigidity scores of 0. Rigidity was detected during the use of an activation maneuver in one control. PD participants had rigidity scores between 0-3 (mean: 1, SD: 1). People with PD had significantly larger clinical rigidity scores than controls (*z* = 37, *p =* 0.001). Leg rigidity was positively correlated with JND threshold (ρ = 0.50, *p =* 0.047), meaning that PD participants with a higher JND threshold tended to be higher rigidity scores (see **Figure 3B**). Uncertainty area did not correlate with the clinical assessment of lower extremity rigidity (*p* > 0.05, **Figure 3C**).

**Figure 3.**
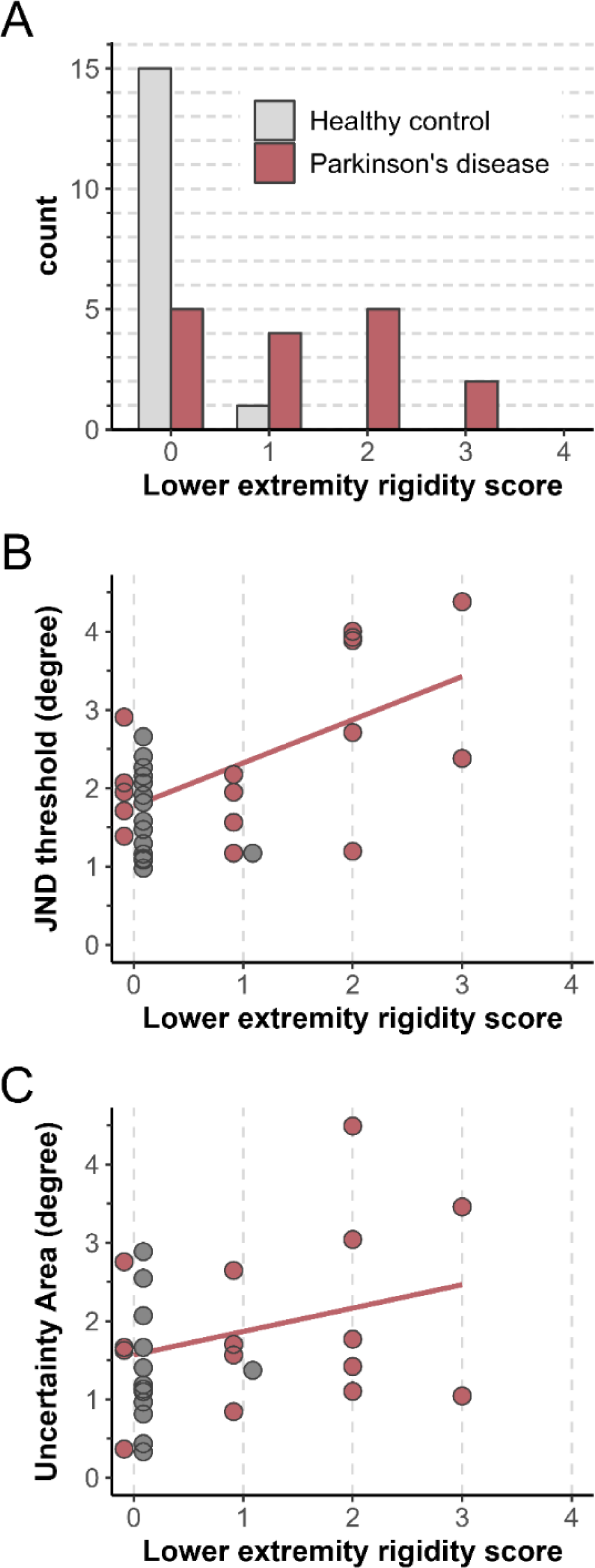
**(A)** The histogram illustrates the distribution of leg rigidity scores between people with Parkinson’s disease (PD, red) and healthy controls (HC, grey). Each bar represents a range of rigidity scores with the height of the bar corresponding to the number of participants falling within that score. Spearman correlation of rigidity score against measures of position sense acuity **(B)** JND position sense threshold and **(C)** UA position sense values in controls and people with Parkinson’s disease. JND position sense threshold significantly increased with increasing rigidity scores in both the PD group (solid red line, *p* = 0.47), meaning that as rigidity increases as proprioceptive bias declines.

### Effects of Disease Duration, Disease Severity and Medication on Conscious and Unconscious Proprioceptive Outcome Measures

Disease duration was moderately correlated with JND threshold (*r* = 0.52, *p* = 0.039) and the clinical assessment of rigidity (ρ = 0.57, *p* = 0.020). JND threshold also correlated moderately with levodopa equivalent dosage (*r* = 0.54, *p* = 0.03). There were no other significant correlations. UPDRS motor score did not yield significant correlations with any outcome measure.

## DISCUSSION

This study examined ankle position sense acuity in PD and the relationship to muscle rigidity. This study applied a psychophysical paradigm that yielded objective and precise measures of ankle position sense bias and precision in people with PD. The main findings are: First, ankle position sense JND threshold and UA are impaired in people with PD. Second, the clinical assessment of rigidity scaled with position sense JND threshold, suggesting that proprioception declines with increases in rigidity. Third, both JND threshold and the clinical impression of rigidity increased with increasing disease duration.

### Parkinson’s disease associated changes in proprioception

There is solid empirical evidence showing that PD may degrade the acuity of all modalities of conscious proprioceptive function – the sense of heaviness, motion and position – in the upper extremities (for a review, see Konczak et al., 2009). Despite its critical role in balance and gait control, only sparse and inconclusive empirical data on lower extremity proprioceptive function are available.

Two previous studies obtained data on position sense acuity at the knee and ankle and both have shown deficits in the Parkinsonian group relative to controls (Ribeiro Artigas et al., 2016; Teasdale et al., 2017). The study by Riberio Artigas and colleagues (2016) found active knee repositioning errors ∼2 times larger in people with PD than controls, which is similar to the magnitude of the deficit seen at the forearm in PD (Maschke et al., 2003). However, the active position matching paradigm applied in these studies did not dissociate the known motor deficits in PD from possible proprioceptive dysfunction. The present study assessed the proprioceptive sense isolated from the motor system.

The proprioceptive outcomes in the control cohort are comparable to those found previously (Sertic et al., 2023). At the group level, people with PD exhibited elevated thresholds of proprioceptive perception. It has been previously reported that 27 – 66% of people with PD demonstrate perception outside of the control range (Konczak et al., 2008; Maschke et al., 2003). In the present study, 38% of people with PD showed elevated JND thresholds and 20% had elevated UA values above the control range, resulting in a total of 44% of participants with a deficit in at least one of the position sense outcomes. To appreciate these data, consider that perceptual bias between two stimuli is proportional to the stimulus intensity – a relationship known as Weber’s law (Bullough et al., 2023). As a proportion of the 15° amplitude reference position, control JND threshold of 1.5° yields a threshold that is 10% of the reference. In contrast, people with PD with elevated thresholds were more than double that of controls (i.e., 24% of the reference).

### Relationship between position sense perception and muscle rigidity in Parkinson’s disease

Proprioceptive dysfunction in PD is not attributable to peripheral nervous system disease in the early disease stage. Muscle spindle afferents to the primary somatosensory cortex and the monosynaptic stretch reflex have normal latencies and magnitudes, which is consistent with normal mechanoreceptor and afferent sensory pathway function (Asci et al., 2023; Lee & Tatton, 1975; Seiss et al., 2003; Tatton & Lee, 1975). In later stages of the disease, there is histological evidence showing PD-related muscle spindle abnormalities (e.g., enlargements in the diffuse endings) that are unrelated to normal aging (Saito et al., 1978). Rather than peripheral mechanoreceptor dysfunction, evidence suggests impairments in central processing of proprioceptive information as the cause of proprioceptive dysfunction in PD (Khudados et al., 1999; Rickards & Cody, 1997; Seiss et al., 2003).

Parkinsonian rigidity is the velocity-dependent upregulation of muscle tone during passive stretch (Asci et al., 2023). A stretch response from the stretched muscle implies that proprioceptive afferents play a role in the generation of rigidity. An exaggerated transcranial long-latency stretch reflex response from the stretched muscle has been found to correlate to the clinical impression of rigidity (Berardelli et al., 1983; Rothwell et al., 1983; Tatton & Lee, 1975). We here report that the impairment of proprioceptive function is related to the clinical impression of rigidity, suggesting that proprioceptive processing deficits are related to both position sense perception and muscle rigidity.

Proprioceptive processing deficits in the present sample were not reflected by tremor-dominant or akinetic rigid subtypes. There were an approximately equal number of participants in each subtype with impairments in position sense and rigidity. Contrary to these findings, Ribeiro Artigas and colleagues (2016) show that people with tremor dominant subtype had greater proprioceptive deficits than people with akinetic rigid subtype (Ribeiro Artigas et al., 2016). However, they employed an active knee joint repositioning task requiring the participant to reposition the knee to the remembered position. Tremor during active movements may yield larger errors in the proprioceptive task, potentially leading to inflated errors.

### Relationship of disease duration and anti-parkinsonian medication to sensory function

Expectedly, severity of rigidity increased with disease duration. It was also found that position sense thresholds increased with the length of disease, corroborating earlier findings (Elangovan et al., 2018; Maschke et al., 2003). In our sample, people with shorter disease duration had JND thresholds comparable to controls whereas people with longer disease durations tended to have elevated thresholds. Braak’s stage 3 is considered the point in disease progression when the pathology appears in the midbrain but the cortex is still mostly uninvolved (Braak et al., 2006). Upon entering stage 4, enough neurons of the substantia nigra pars compacta have degenerated to give rise to the clinically recognizable motor phase of the disease. The basal ganglia has proprioceptive receptive fields, yet somatosensation may be preserved until Lewy bodies and α-synucleophathies affect cortical regions in disease stages 5 and 6 (Rodriguez-Oroz et al., 2001). However, this is only speculative as we do not have data on lesion locations in our participant sample.

Somatosensory function as it relates to the total MDS-UPDRS motor score is unclear as some studies indicate worsening function with increased disease severity whereas others show no relationship (Elangovan et al., 2018; Konczak et al., 2007, 2008; Maschke et al., 2003, 2006). The present study found that neither position sense acuity measure correlated with the total MDS-UPDRS motor score. It is important to note that our sample consisted of mild-to-moderate PD with the most severe participant scoring 52 on the MDS-UPDRS III (maximum score 132). At present, it is unclear how proprioceptive function may present at later stages of PD as it is difficult to recruit this patient demographic to the lab.

Our study sheds light on the nuanced relationship between anti-parkinsonian medication dosage and proprioceptive function. Contrary to most prior reports predominantly conducted in the ON medication state, which largely found no significant relationship between L-dopa dosage and somatosensory function, our data indicate a decline in proprioceptive abilities with increasing medication dosage (Elangovan et al., 2018; Konczak et al., 2007, 2012; Maschke et al., 2003, 2006). This disparity suggests that previous studies may have overlooked the potential confounding effects of medication on somatosensory function. While there is conflicting evidence on whether L-dopa has an influence on the proprioceptive sense, the one study with a similar design as employed by this study found that anti-parkinsonian medication improved proprioceptive outcomes by ∼15% (Li et al., 2010; O’Suilleabhain, 2001; Wright et al., 2010). Our results, derived from participants washed out of medication, unveil the true disease state of proprioceptive function and hint at the possibility of a therapeutic role for medication in managing proprioceptive deficits in PD.

### Limitations

The sample population consisted of individuals with PD who were highly active and predominately exhibited mild disease severity. Consequently, the generalizability of our results to individuals with more advanced stages of the disease are limited.

## CONCLUSIONS

We employed a psychophysical paradigm to assess ankle position sense acuity and clinically assessed rigidity in people with PD and age-matched control counterparts. There were 44% of participants with PD with abnormalities in either JND threshold or UA, or both. These findings complement previous research identifying upper extremity proprioceptive deficit. It provides evidence that the impairment also generalizes the lower extremities. Given that ankle proprioceptive deficits impair balance and gait, and given that proprioception can be trained in PD, future research may explore an ankle proprioceptive training task to improve static and dynamic balance (Elangovan et al., 2018). The present study also associated impairments in JND threshold with elevated muscle rigidity. This implies that impairments in proprioceptive processing contribute to both proprioceptive dysfunction and rigidity in PD.

## Data Availability

All data produced in the present study are available upon reasonable request to the authors.

## ACKNOWLEDGEMENTS

We thank all the people who volunteered for this study. We thank Dr. Paul Tuite and Joshua de Kam for their assistance in recruitment of people with Parkinson’s disease. This research was supported by a grant from the North American Society for the Psychology of Sport and Physical Activity and an award from the Center for Clinical Movement Science at the University of Minnesota to JVL Sertic. Additional support came from the UMN Graduate School Doctoral Dissertation Fellowship to JVL Sertic.

## Author Contributions

JVL Sertic, J Konczak, and CD MacKinnon contributed to the study conception and design. Data collection was performed by JVL Sertic and J Kang. Data analyses were performed by JVL Sertic. The first draft of the manuscript was written by JVL Sertic and all authors commented on subsequent versions of the manuscript. All authors read and approved the final manuscript.

